# Semaglutide as an Adjunctive Therapy to Standard Management for Idiopathic Intracranial Hypertension: A Real-World Data-Based Retrospective Analysis

**DOI:** 10.1101/2024.11.12.24317197

**Authors:** Ahmed Y. Azzam, Muhammed Amir Essibayi, Dhrumil Vaishnav, Mohammed A. Azab, Mahmoud M. Morsy, Osman Elamin, Ahmed Saad Al Zomia, Hammam A. Alotaibi, Ahmed Alamoud, Adham A. Mohamed, Omar S. Ahmed, Adam Elswedy, Oday Atallah, Hana J. Abukhadijah, Adam A. Dmytriw, David J. Altschul

**Author notes:** **Corresponding Author:** Hana J. Abukhadijah, Medical Research Center, Hamad Medical Corporation, Doha Qatar, P.O. Box 3050.

## Abstract

**Background:** Idiopathic intracranial hypertension (IIH) is a neurological disorder characterized by elevated intracranial pressure, predominantly affecting young women with obesity. This study evaluates the effectiveness of semaglutide as an adjunctive therapy to standard IIH management using real-world data.

**Methods:** We conducted a retrospective cohort analysis comparing IIH patients receiving semaglutide plus standard therapy versus standard therapy alone. After propensity score matching, we analyzed 635 patients in each cohort. Primary outcomes included papilledema, headache manifestations, visual disturbances, and refractory disease status at 3-months, 6-months, 12-months, and 24-months. Secondary outcomes included BMI changes.

**Results:** Semaglutide demonstrated significant improvements across all outcomes. At three months, the treatment group showed reduced risks of visual disturbances (RR 0.28, 95% CI 0.179-0.440, p=0.0001), papilledema (RR 0.366, 95% CI 0.260-0.515, p=0.0001), and headache (RR 0.578, 95% CI 0.502-0.665, p=0.0001). These benefits persisted through 24 months. Refractory disease risk was reduced by 40% at three months (RR 0.6, 95% CI 0.520-0.692, p=0.0001). The semaglutide group showed progressive BMI reduction, with a baseline-adjusted difference of -1.38 kg/m^2^ (95% CI [-1.671, -1.089], p<0.0001) at 24 months.

**Conclusions:** Semaglutide as an adjunctive therapy demonstrates significant and sustained improvements in IIH-related outcomes, including visual disturbances, papilledema, and headache symptoms. These findings suggest semaglutide may be a valuable addition to standard IIH management protocols, particularly for patients with refractory disease.

## 1. Introduction

Idiopathic intracranial hypertension (IIH) is a neurological disorder characterized by elevated intracranial pressure (ICP) without identifiable cause, predominantly affecting young women with obesity. The condition manifests through chronic headaches, papilledema, and potential permanent vision loss if left untreated. Recent epidemiological data indicate rising IIH incidence paralleling global obesity trends, with significant healthcare burden and reduced quality of life among affected individuals. Current management approaches include medical therapy with acetazolamide, topiramate, and diuretics as first-line treatments, while surgical interventions such as cerebrospinal fluid (CSF) shunting, optic nerve sheath fenestration, and venous sinus stenting are reserved for medically refractory cases [1-3]. However, existing therapeutic options have notable limitations. Medical treatments often provide suboptimal ICP reduction and are frequently limited by poor tolerability. Acetazolamide, the most commonly prescribed medication, is associated with significant side effects including cognitive impairment, with up to 48% of patients discontinuing therapy within three months. Surgical interventions, while effective, carry procedural risks and may require revision surgeries [4, 5].

Glucagon-like peptide-1 receptor agonists (GLP-1 RAs) have emerged as a promising therapeutic avenue in IIH management. These agents, originally developed for type 2 diabetes and obesity treatment, work through multiple mechanisms including appetite suppression via hypothalamic signaling and direct effects on CSF production through GLP-1 receptors expressed in the choroid plexus. Recent evidence from the IIH:Pressure trial demonstrated that exenatide, a GLP-1 RA, significantly reduced ICP by 5.7 cmCSF at 2.5 hours, with sustained effects at 12 weeks (5.6 cmCSF reduction). Additionally, the recent VIIH database analysis showed significant improvements in headache outcomes with GLP-1 RA therapy, with 76.9% of patients achieving ≥50% reduction in monthly headache days [6-8]. Our study aims to evaluate the real-world effectiveness of semaglutide as an adjunctive therapy to standard IIH management using the TriNetX database. This retrospective cohort analysis leverages a large-scale, multi-institutional database to provide robust evidence regarding semaglutide’s role in IIH treatment.

The study’s strengths lie in its substantial sample size, real-world setting, and comprehensive assessment of clinical outcomes across multiple institutions, offering valuable insights into the practical application of semaglutide in routine clinical care for IIH patients [9].

## 2. Methods

### 2.1. Study Design and Data Source

We conducted a retrospective cohort analysis utilizing the TriNetX Global Health Research Network (TriNetX, Cambridge, MA, USA), a federated real-time platform integrating electronic health records from approximately 160 healthcare organizations worldwide [10]. The network encompasses around 197 million patient records across multiple countries, including the United States as the predominant source, along with healthcare data from Australia, Belgium, Brazil, Bulgaria, Estonia, France, Georgia, Germany, Ghana, Israel, Italy, Japan, Lithuania, Malaysia, Poland, Singapore, Spain, Taiwan, United Arab Emirates, and the United Kingdom. Our study leveraged data through October 2024, representing the complete available timeframe in the dataset (**Figure 1**).

**Figure 1.**
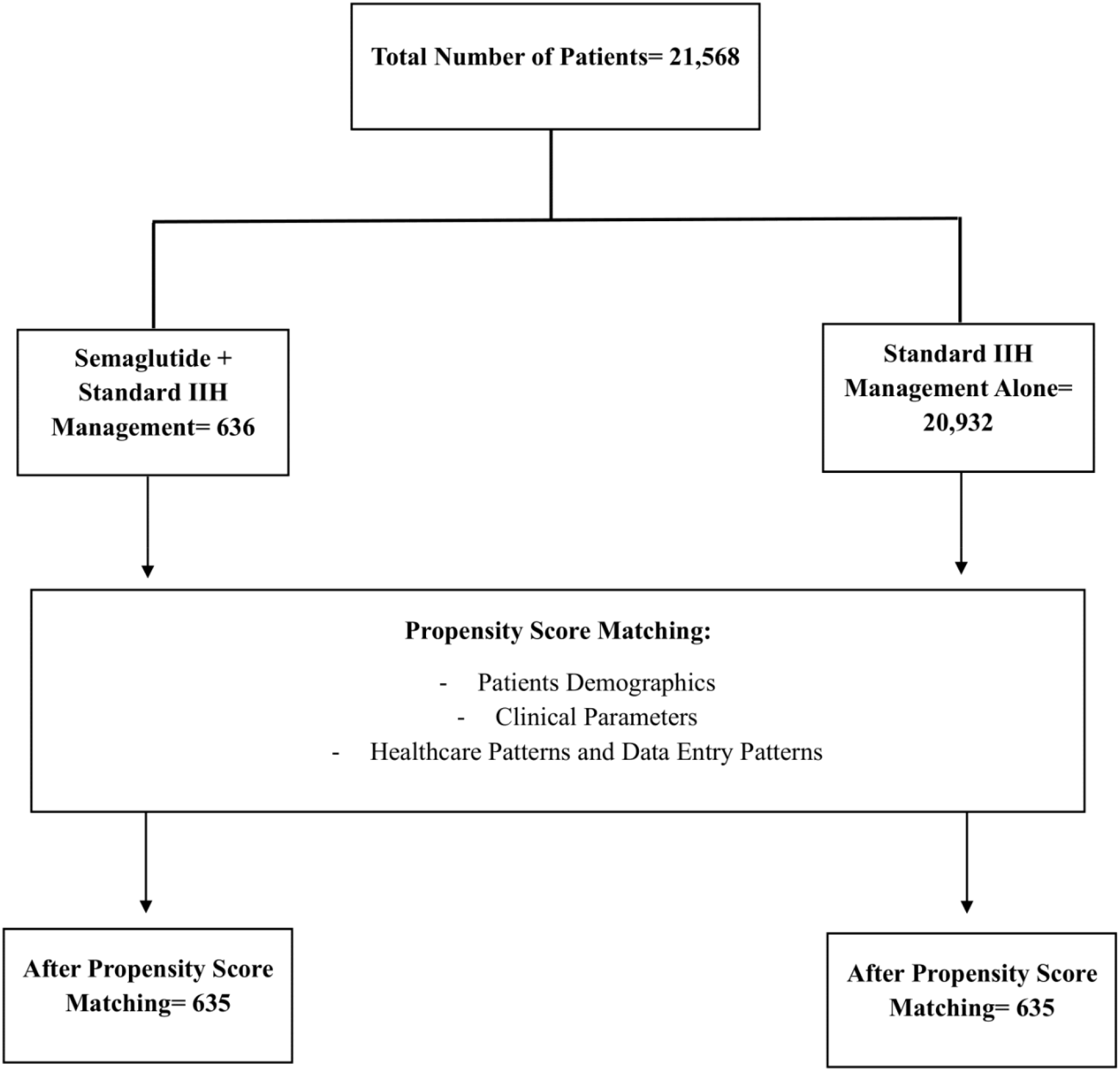
Patients Selection Flow Diagram.

The TriNetX platform provides comprehensive patient-level information including demographics, diagnoses, treatments, procedures, and clinical outcomes, coded using standardized medical classification systems such as the International Classification of Diseases, Tenth Revision (ICD-10) and Current Procedural Terminology (CPT) codes. All data within the network are automatically standardized and harmonized to enable consistent analysis across different healthcare organizations and geographical regions. The platform ensures HIPAA compliance through automatic de-identification of patient data while maintaining the integrity of longitudinal records for research purposes.

### 2.2. Study Population and Cohort Definition

We systematically identified patients with IIH using the International Classification of Diseases, Tenth Revision, Clinical Modification (ICD-10-CM) code G93.2 (Benign intracranial hypertension). The inclusion criteria were rigorously defined to include age ≥18 years at the time of diagnosis, confirmed IIH diagnosis with at least one documented clinical encounter, active engagement in standard medical therapy as evidenced by prescription records, and availability of baseline clinical measurements including BMI documentation.

The semaglutide cohort was defined through prescription records indicating semaglutide initiation (identified through RxNorm concept unique identifiers) as an adjunctive therapy to standard management protocols. The control cohort comprised patients receiving standard therapy alone, typically including acetazolamide, topiramate, or other diuretics, in addition to weight loss / management approaches without exposure to GLP-1 receptor agonists.

We implemented a specific exclusion criterion to ensure data integrity. These included secondary causes of intracranial hypertension identified through relevant ICD-10 codes, including cerebral venous thrombosis, medication-induced intracranial hypertension, or structural central nervous system abnormalities. Additional exclusion criteria encompassed concurrent or previous use of other GLP-1 receptor agonists within six months of cohort entry, pregnancy or immediate post-partum status, absence of baseline BMI measurements, and follow-up period less than three months. Cases with missing critical data elements were systematically excluded to maintain analytical rigor.

### 2.3. Propensity Score Matching Methodology

To mitigate selection bias and establish comparable cohorts, we implemented a 1:1 propensity score matching protocol utilizing TriNetX’s built-in analytics features. The propensity score model incorporated multiple variables including demographic characteristics such as age at diagnosis, sex, race (categorized as White, Black or African American, Asian, and Other), and ethnicity (Hispanic or Latino, Non-Hispanic or Latino). Clinical parameters included baseline BMI, comorbidity profiles including endocrine, musculoskeletal, and eye disorders, pre-existing medication usage, and healthcare utilization patterns in the year preceding cohort entry.

The matching process employed a greedy nearest-neighbor algorithm with a caliper width of 0.2 standard deviations of the logit of the propensity score. This rigorous matching methodology refined our initial sample of 636 semaglutide-treated patients and 20,932 controls to balanced cohorts of 635 patients each, achieving optimal covariate balance between groups.

### 2.4. Outcome Measures and Assessment

Primary outcomes were systematically tracked through structured query logic within the TriNetX platform at predetermined intervals of three-months, six-months, 12-months, and 24-months post-cohort entry. The primary outcome measures included papilledema (ICD-10 code H47.1), headache manifestations (ICD-10 codes G44.-), and visual disturbances or blindness (ICD-10 codes H53.-and H54.-). Refractory IIH was defined as either persistence of symptoms despite maximum medical therapy, progression to surgical intervention (identified through relevant CPT codes), or requirement for CSF diversion procedures or optic nerve sheath fenestration.

Secondary outcomes focused on BMI trajectories, with measurements extracted directly from structured clinical data fields at each follow-up timepoint. We calculated both absolute BMI changes and baseline-adjusted differences to account for initial between-group disparities.

### 2.5. Statistical Analysis Framework

All analyses were performed using TriNetX’s analytical tools, which employ standardized statistical methodologies. For primary outcomes, we calculated relative risks (RR) with corresponding 95% confidence intervals, absolute risk differences between groups, and number needed to treat (NNT) where appropriate. Continuous variables were expressed as means ± standard deviations, with between-group comparisons conducted using Student’s t-tests.

Categorical variables were analyzed using chi-square tests or Fisher’s exact tests where appropriate. For BMI analysis, we computed both absolute and baseline-adjusted differences between groups, with temporal trends assessed through longitudinal analysis features. Statistical significance was consistently defined as p<0.05, with exact p-values reported where available. Multiple comparison adjustments were applied using the Benjamini-Hochberg procedure to control the false discovery rate.

## 3. Results

In our propensity score-matched analysis, we evaluated baseline characteristics between semaglutide-treated and control cohorts (**Table 1**). Our initial sample comprised 636 patients in the semaglutide group and 20,932 in the control group, which was subsequently matched to achieve balanced cohorts of 635 patients each. The demographic composition revealed a predominantly female population (93.1%) in both matched groups. Mean age was comparable between matched cohorts (37.6 ± 9.01 vs 37.1 ± 9.4 years, p=0.3403), though we observed a persistent significant difference in body mass index (BMI), (41.5 ± 8.68 vs 37.2 ± 8.9 kg/m^2^, p<0.0001). Our matched cohorts demonstrated significant balance across racial and ethnic distributions, with White patients constituting the majority (65.4% vs 70.2%, p=0.0627), followed by Black or African American (19.5% vs 17.2%, p=0.2768) and Hispanic or Latino patients (9.0% vs 6.6%, p=0.1164). Notably, we achieved a balance in comorbidity profiles post-matching, with comparable rates of endocrine (88.5% vs 89.3%, p=0.6552), musculoskeletal (69.4% vs 68.3%, p=0.6713), and eye disorders (69.3% vs 67.2%, p=0.4332).

**Table 1.** Baseline Characteristics of Semaglutide Group and Control Group Before and After Propensity Score Matching for Cohorts.

| **Variable** | **Semaglutide Before Matching** | **Control Before Matching** | **P-value Before** | **Semaglutide After Matching** | **Control After Matching** | **P-value After** |
| --- | --- | --- | --- | --- | --- | --- |
| Sample size, n | 636 | 20932 | N/A | 635 | 635 | N/A |
| Age (years), mean ± SD | 37.6 ± 9.01 | 35.6 ± 10 | < 0.0001 | 37.6 ± 9.01 | 37.1 ± 9.4 | 0.3403 |
| BMI, mean ± SD | 41.5 ± 8.68 | 35.4 ± 9.04 | < 0.0001 | 41.5 ± 8.68 | 37.2 ± 8.9 | < 0.0001 |
| Female, n (%) | 591 (93.1%) | 17852 (85.8%) | < 0.0001 | 591 (93.1%) | 591 (93.1%) | 0.999 |
| Male, n (%) | 17 (2.7%) | 2533 (12.2%) | < 0.0001 | 17 (2.7%) | 19 (3.0%) | 0.7352 |
| ***Race/Ethnicity:*** |  |  |  |  |  |  |
| White, n (%) | 415 (65.4%) | 12419 (59.7%) | 0.004 | 415 (65.4%) | 446 (70.2%) | 0.0627 |
| Black or African American, n (%) | 124 (19.5%) | 3990 (19.2%) | 0.8229 | 124 (19.5%) | 109 (17.2%) | 0.2768 |
| Hispanic or Latino, n (%) | 57 (9.0%) | 1930 (9.3%) | 0.7989 | 57 (9.0%) | 42 (6.6%) | 0.1164 |
| Asian, n (%) | 10 (1.6%) | 299 (1.4%) | 0.7737 | 10 (1.6%) | 10 (1.6%) | 0.999 |
| ***Comorbidities:*** |  |  |  |  |  |  |
| Endocrine disorders, n (%) | 562 (88.5%) | 8676 (41.7%) | < 0.0001 | 562 (88.5%) | 567 (89.3%) | 0.6552 |
| Musculoskeletal disorders, n (%) | 441 (69.4%) | 6774 (32.6%) | < 0.0001 | 441 (69.4%) | 434 (68.3%) | 0.6713 |
| Eye disorders, n (%) | 440 (69.3%) | 11620 (55.8%) | < 0.0001 | 440 (69.3%) | 427 (67.2%) | 0.4332 |
| Digestive system disorders, n (%) | 365 (57.5%) | 4984 (23.9%) | < 0.0001 | 365 (57.5%) | 357 (56.2%) | 0.6504 |
| Circulatory system disorders, n (%) | 175 (27.6%) | 3501 (16.8%) | < 0.0001 | 175 (27.6%) | 176 (27.7%) | 0.95 |

### 3.1. Symptoms and Outcomes Analysis

Our analysis demonstrated significant therapeutic benefits of semaglutide as an adjunctive therapy across all measured outcomes in patients with IIH (**Table 2**). We observed statistically significant improvements in visual outcomes, with the semaglutide group showing a 72% reduction in risk of visual disturbances or blindness at three months (RR 0.28, 95% CI 0.179-0.440, p=0.0001). This protective effect persisted throughout the follow-up period, with a sustained risk reduction of 62% at 24-months (RR 0.38, 95% CI 0.279-0.518, p=0.0001).

**Table 2.** Outcomes Relative Risk For Semaglutide Group Compared to Control Group.

| **Outcome** | **Follow-up Timepoint** | **Risk Ratio** | **95% Confidence Interval** | **P-value** |
| --- | --- | --- | --- | --- |
| Papilledema | 3-months | 0.366 | (0.260, 0.515) | 0.0001 |
|  | 6-months | 0.415 | (0.308, 0.559) | 0.0001 |
|  | 12-months | 0.486 | (0.374, 0.632) | 0.0001 |
|  | 24-months | 0.497 | (0.386, 0.639) | 0.0001 |
| Headache | 3-months | 0.578 | (0.502, 0.665) | 0.0001 |
|  | 6-months | 0.672 | (0.600, 0.753) | 0.0001 |
|  | 12-months | 0.705 | (0.638, 0.779) | 0.0001 |
|  | 24-months | 0.718 | (0.653, 0.788) | 0.0001 |
| Refractory IIH | 3-months | 0.6 | (0.520, 0.692) | 0.0001 |
|  | 6-months | 0.702 | (0.625, 0.789) | 0.0001 |
|  | 12-months | 0.735 | (0.663, 0.815) | 0.0001 |
|  | 24-months | 0.751 | (0.682, 0.827) | 0.0001 |
| Visual Disturbances or Blindness | 3-months | 0.28 | (0.179, 0.440) | 0.0001 |
|  | 6-months | 0.303 | (0.204, 0.449) | 0.0001 |
|  | 12-months | 0.363 | (0.258, 0.510) | 0.0001 |
|  | 24-months | 0.38 | (0.279, 0.518) | 0.0001 |

Papilledema has also showed significant improvement in the semaglutide group over the control group. Our results demonstrated a 63.4% reduction in risk at three months (RR 0.366, 95% CI 0.260-0.515, p=0.0001), with sustained benefit at 24-months showing a 50.3% risk reduction (RR 0.497, 95% CI 0.386-0.639, p=0.0001). This finding carries particular clinical significance given the potential for permanent visual loss in untreated papilledema. In addition to that, we found that headache symptoms, often the most debilitating complaint among IIH patients, responded favorably to semaglutide therapy. The intervention group demonstrated a 42.2% reduction in headache risk at three months (RR 0.578, 95% CI 0.502-0.665, p=0.0001), with continued benefit showing a 28.2% risk reduction at 24-months (RR 0.718, 95% CI 0.653-0.788, p=0.0001).

Our results revealed a good impact on refractory IIH cases, traditionally the most challenging to manage, which failed the current standard management approaches alone. The semaglutide group showed a 40% reduction in risk of refractory disease at three months (RR 0.6, 95% CI 0.520-0.692, p=0.0001), maintaining a 24.9% risk reduction at 24-months (RR 0.751, 95% CI 0.682-0.827, p=0.0001). The consistency of risk reductions across all outcome measures and time points, coupled with tight confidence intervals, strengthens the validity of our findings. While we observed some attenuation of effect sizes over the 24-month follow-up period, the maintenance of statistically significant risk reductions suggests durable therapeutic benefit with semaglutide as an adjunctive therapy in IIH management.

The risk difference longitudinal analysis revealed a distinctive temporal pattern in treatment response across all outcomes. We observed that the maximum therapeutic divergence between semaglutide and control groups consistently occurred at the six-month follow-up timepoint, followed by a slight convergence that stabilized by 12-months as demonstrated in **Figure 2**. This pattern was particularly evident in visual disturbances and papilledema, where the risk differential reached its nadir at six months (approximately 11% versus 20% for papilledema, and 5% versus 15% for visual disturbances in semaglutide versus control groups, respectively). The subsequent modest convergence in risk trajectories after six months suggests an initial period of maximal therapeutic efficacy, followed by a sustained but slightly attenuated long-term benefit. The absolute risk reduction remained clinically significant even at 24 months, with the semaglutide group maintaining consistently lower risk profiles across all outcomes.

**Figure 2.**
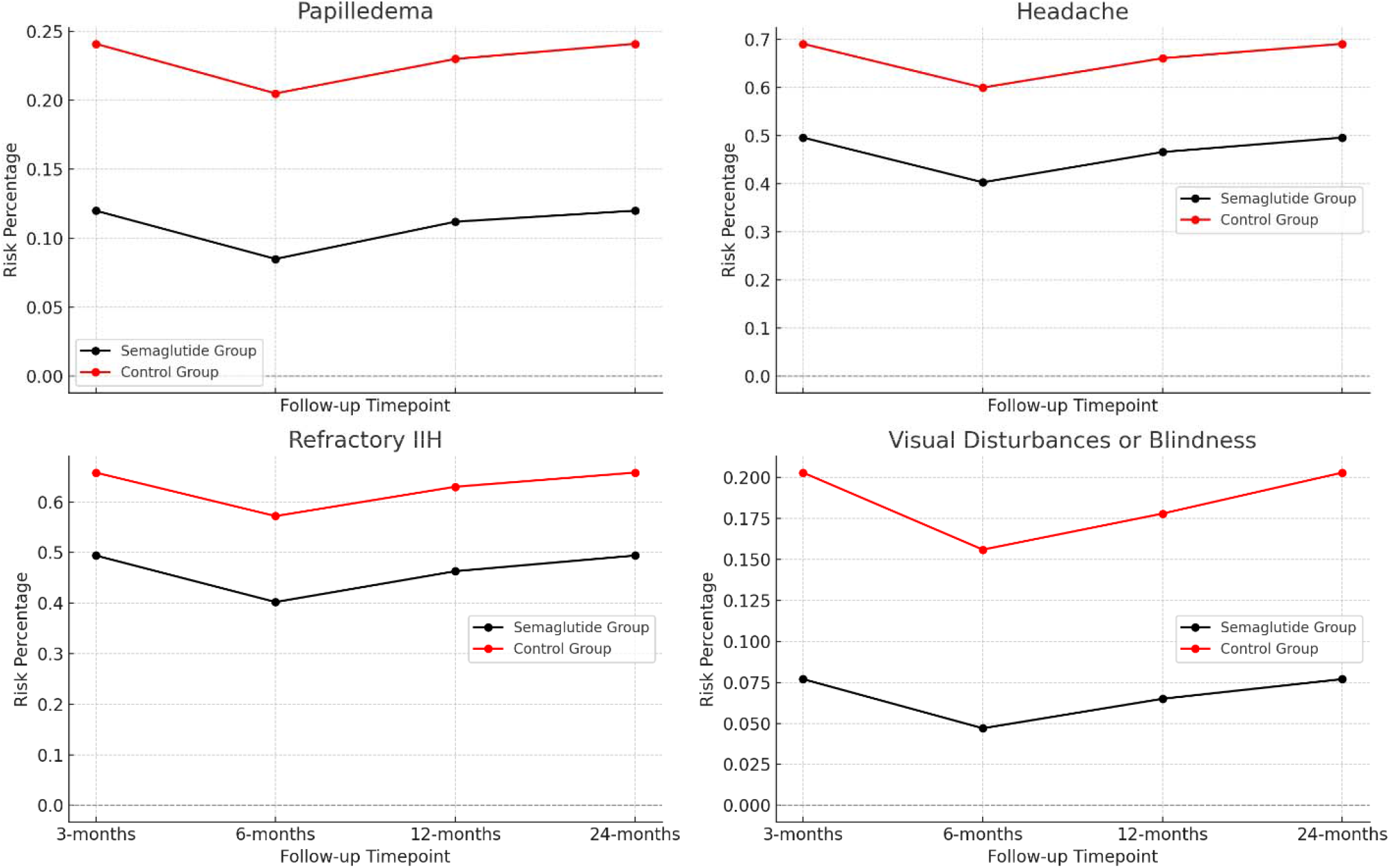
Temporal Risk Difference For Key Clinical Outcomes.

### 3.2. BMI Longitudinal Analysis

We performed a timepoint based longitudinal analysis of BMI trends which revealed significant differences between the semaglutide and control groups throughout the 24-month follow-up period (**Table 3**). At baseline, the semaglutide group presented with a higher mean BMI (41.5 ± 8.68 kg/m^2^) compared to the control group (37.2 ± 8.9 kg/m^2^). Despite this initial disparity, we observed a consistent pattern of BMI reduction in the semaglutide group over time. The baseline-adjusted BMI differences demonstrated a progressive and statistically significant decline in the semaglutide group. The initial modest reduction at three months (-0.357 kg/m^2^, 95% CI [-0.687, -0.027], p=0.03) evolved into more decrements at subsequent time points. By six months, the baseline-adjusted BMI difference had increased to -0.874 kg/m^2^ (95% CI [-1.183, -0.565], p<0.0001), further progressing to -1.141 kg/m^2^ (95% CI [-1.437, -0.845], p<0.0001) at 12-months. The maximal weight loss difference effect was observed at 24-months, with a baseline-adjusted BMI difference of -1.38 kg/m^2^ (95% CI [-1.671, -1.089], p<0.0001).

**Table 3.**
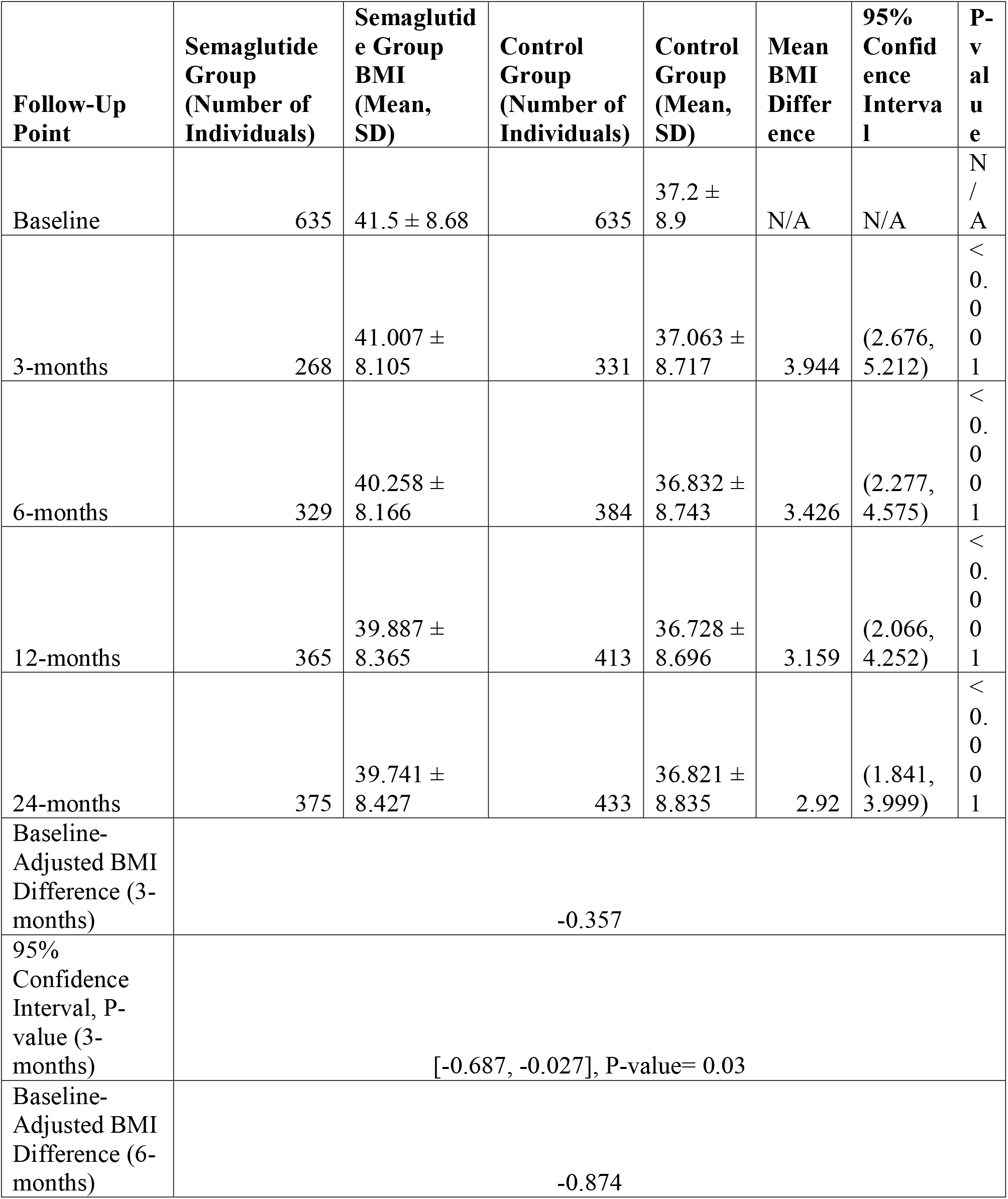
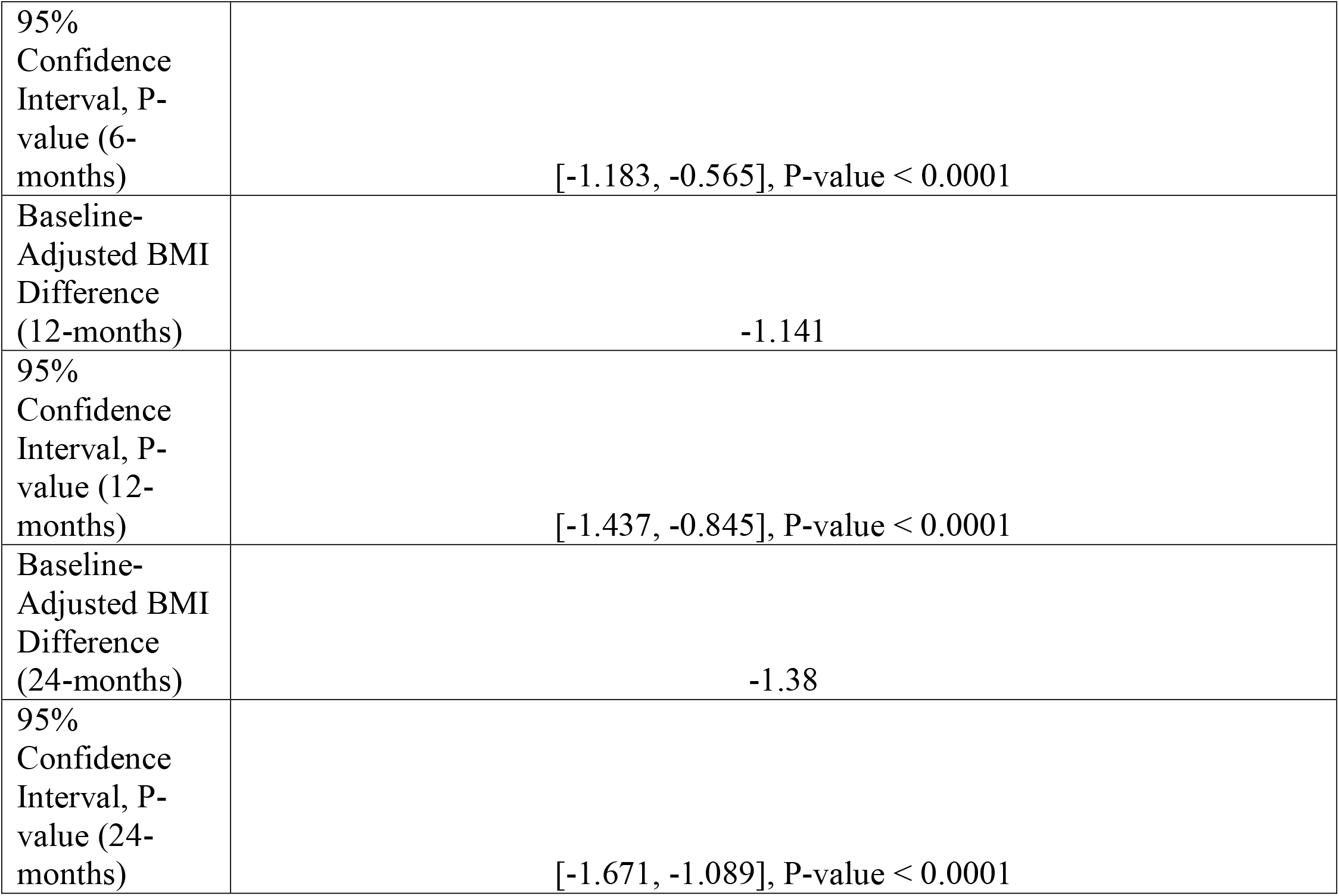
BMI Differences Over Follow-Time Point Comparison Between Semaglutide Group vs. Control Group.

We noted differential patterns in follow-up participation between groups, with varying numbers of individuals at each time point. The semaglutide group showed consistent engagement, progressing from 268 individuals at three months to 375 at 24 months, while the control group maintained slightly higher follow-up numbers, ranging from 331 to 433 participants. Despite these variations in follow-up patterns, the absolute BMI differences between groups remained statistically significant throughout the study period, ranging from 3.944 kg/m^2^ (95% CI [2.676, 5.212], p<0.001) at three months to 2.92 kg/m^2^ (95% CI [1.841, 3.999], p<0.001) at 24-months. This sustained difference, coupled with the progressive baseline-adjusted reductions, suggests a durable effect of semaglutide on weight management in our IIH cohort over the standard management alone.

## 4. Discussion

The emergence of GLP-1 RAs represents a promising therapeutic avenue for IIH, extending beyond their established role in weight management. Recent evidence has demonstrated that GLP-1 receptors are expressed in the choroid plexus, suggesting a direct mechanism for modulating CSF dynamics independent of weight loss effects [6, 7]. This dual mechanism of action positions GLP-1 RAs as particularly relevant therapeutic agents for IIH management [11-14].

Our study represents the first large-scale, real-world analysis of semaglutide’s effectiveness as an adjunctive therapy in IIH management using the TriNetX Global Health Research Network [10]. The study’s strengths lie in its substantial sample size, comprehensive outcome assessment across multiple institutions, and real-world setting, offering insights into the practical application of semaglutide in routine clinical care. By leveraging data from international healthcare organizations worldwide, our analysis provides robust evidence regarding semaglutide’s role in IIH treatment while maintaining methodological rigor through propensity score matching to minimize selection bias [15-18].

The findings demonstrate significant improvements across multiple outcome measures in the semaglutide-treated cohort. Most notably, we observed a 72% reduction in risk of visual disturbances or blindness at three months (RR 0.28, 95% CI 0.179-0.440, p=0.0001), with sustained benefits at 24 months. This aligns with recent findings from the IIH:Pressure trial, which demonstrated significant reductions in intracranial pressure with GLP-1 RA therapy [6, 8]. However, our study extends these findings by demonstrating sustained benefits in a larger, real-world population over a longer follow-up period. Particularly noteworthy is the significant improvement in papilledema outcomes, with a 63.4% reduction in risk at three months (RR 0.366, 95% CI 0.260-0.515, p=0.0001). This finding is especially relevant given that the IIH:Pressure trial, while demonstrating reduced intracranial pressure with exenatide, was not powered to evaluate long-term visual outcomes [6, 8]. Our results suggest that GLP-1 RA therapy may provide sustained protection against one of IIH’s most concerning complications.

The headache outcomes in our study parallel recent findings from the VIIH database analysis, which showed significant improvements with GLP-1 RA therapy [8]. Our observation of a 42.2% reduction in headache risk at three months (RR 0.578, 95% CI 0.502-0.665, p=0.0001) provides additional evidence supporting the role of GLP-1 RAs in managing this debilitating aspect of IIH.

Our findings regarding refractory IIH cases are particularly encouraging, showing a 40% reduction in risk at three months. This compares favorably with previous studies of medical therapy in refractory cases [1, 19] and suggests that GLP-1 RAs might offer an alternative to surgical intervention in selected cases with proper evaluation and consideration [20]. The sustained benefits observed over 24 months also address an important gap in the literature regarding long-term medical management options for refractory IIH [1, 19, 20].

Several limitations of our study warrant consideration. First, the retrospective nature of the analysis introduces potential selection bias, although we attempted to minimize this through propensity score matching. Second, the TriNetX platform’s limitations prevented detailed analysis of semaglutide dosing regimens and administration routes, which could provide valuable insights into optimal therapeutic strategies. Additionally, we were unable to stratify outcomes based on the timing of semaglutide initiation relative to IIH diagnosis, which might influence treatment effectiveness. The impact of different semaglutide formulations (oral versus injectable) and dosing frequencies on outcomes remains an important area for future research. Recent evidence suggests that higher doses of GLP-1 RAs may provide additional benefits in weight management [21-24], but the optimal dosing strategy for IIH management remains unclear.

Furthermore, while our study demonstrates sustained benefits over 24 months, longer-term follow-up studies are needed to evaluate the durability of treatment effects and potential development of treatment resistance.

Our findings have important implications for future research and clinical trials directions. Prospective randomized controlled trials should evaluate the optimal timing of GLP-1 RA initiation in IIH management, potentially as first-line therapy rather than as an adjunctive treatment. Additionally, studies comparing different GLP-1 RAs head-to-head in IIH management could help identify the most effective agents within this class. The potential synergistic effects of combining GLP-1 RAs with other IIH treatments, particularly acetazolamide, also warrants investigation, as our results suggest potential benefits of combination therapy.

## Declarations

### IRB Approval

Waived.

### bold>Ethical Approvals

Waived.

### Consent for Participation

N/A.

## Acknowledgements

Open access funding is provided by the Qatar National Library.

## Data Availability Statement

All used data is available within TriNetX database platform.

## LLM Statement

We have employed an advanced Large Language Model (LLM) to enhance and refine the English-language writing. This process focused solely on improving the text’s clarity and style, without generating or adding any new information to the content.

## Author Contributions: Conceptualization

Ahmed Y. Azzam, Hana J. Abukhadijah, and David J. Altschul **Methodology:** Ahmed Y. Azzam, Muhammed Amir Essibayi, and Dhrumil Vaishnav **Data Collection and Analysis:** Mohammed A. Azab, Mahmoud M. Morsy, Osman Elamin, Ahmed Saad Al Zomia, Hammam A. Alotaibi, and Ahmed Alamoud **Clinical Investigation:** Adham A. Mohamed, Omar S. Ahmed, and Adam Elswedy **Data Validation:** Oday Atallah and Adam A. Dmytriw **Project Administration:** Hana J. Abukhadijah Supervision: David J. Altschul **Writing -Original Draft:** Ahmed Y. Azzam, Muhammed Amir Essibayi, and Dhrumil Vaishnav **Writing - Review & Editing:** All authors. All authors have read and agreed to the published version of the manuscript.

